# Patterns of viruses among the jaundice patients admitted in a tertiary care hospital, Chattogram, Bangladesh

**DOI:** 10.1101/2025.01.15.25320599

**Authors:** Rajat Sanker Roy Biswas, Sk Md Faisal Abdullah, Shemanta Waddadar, Nabil Asgar Chowdhury

## Abstract

**Introduction:** Jaundice is the commonest presenting features of hepatitis and it is a common clinical condition in Bangladesh. As a country of the Asia Pacific region Bangladesh is considered to be a high risk country for developing hepatitis by different hepatotropic viruses. This studyrepresents the patterns and types ofviruses among the patients admitted in a tertiary care hospitals inBangladesh who were presented with jaundice.

**Methods:** Present cross sectional observational study done during a one year study period from July 2022 to June 2023 in a tertiary care hospital, Chattogram, Bangladesh among 114 patients. All patients who had clinical jaundice or having serum bilirubin > 2 mg/dl were recruited in the study. Clinical data were collected from all of them and viral serology were also done by ICT or ELISA. Informed written consent were taken from all patients and permission for the study were taken from ERB of the medical college.

**Results:** Among 114 study subjects, male were 73(64%) and female were 41(36%) and male to female ratio was 1: 0.56. Among all, maximum were in the age range of <20 years and 21- 30 years which were 46(40.4%) and 44(38.6%) respectively. Regarding clinical features, 109(95.6%) had jaundice and 89(78.1%) had fever and 74(64.9%) had nausea and these were three dominant clinical features found. Viral serology study revealed frequency of HAV, HBV, HCV and HEV infection were 43(37.7%), 9(7.9%), 1(0.9%) and 26(22.8%) respectively. There were viral coinfections with (HAV+HEV), (HAV+HBV) and (HBV + HEV) and it were 8(7.0%), 4(3.5%) and 3(2.6%) respectively. Among all 20(17.5%) cases were tested negative for hepatotropic viruses. Again 6(5.3%) had Anti HBc Positivity and 2(1.8%) cases were found coinfected with HIV.

**Conclusions:** In a year round, HAV and HEV are two common viral infection causing jaundice among the locality. HBV had a significant level of infection with low level HCV causing jaundice. Two HIV coinfection is an alarming condition found among the jaundice patients which need further exploration and risk factor evaluations.

## Introduction

Viral hepatitis are common infectious disease in Bangladesh.[1] Hepatotropic viruses like A, B, C, and E are the common viruses responsible for it. But other uncommon viruses also may cause jaundice but these viruses mostly remain undiagnosed. Viral hepatitis constitutes huge burdento the health care delivery system and it has an economic impact in Bangladesh.[1]

Among the different hepatotropic viruses hepatitis E virus is the leading cause of acute hepatitis in this country.[2] Forchronic liver disease, hepatitis B virus is one of the main cause and its prevalenceis 5-6% in our country and next to it is hepatitis C and its prevalence is 1%.[3]

Hepatitis A also prevailing in our country and its frequency increases during rainy season. Epidemic and sporadic outbreaks of acute hepatitis in low-income countries, such as

Bangladesh, are commonly caused by HAV and HEV, which are predominantly transmittedvia the fecal–oral route [4]. The clinical manifestations of HAV infection vary from asymptomatic infection to acute liver failure (ALF), but do not lead to chronic hepatitis. Hepatitis A causes mild-to-severe manifestations, includingfever, malaise, loss of appetite, diarrhea, nausea, abdominal discomfort, dark-colored urine,and jaundice. Clinical manifestations of HAV depend on the age of the patients; mildsymptoms are observed in children and severe infections in adults [5].

These four maintypes of hepatitis viruses are of greatestconcern because of the burden of illness and death they cause and the potential for outbreaks andepidemic spread Bangladesh. In particular, types B and C lead to chronic disease in hundreds of millions ofpeople and, together, are the most common cause of liver cirrhosis and cancer[6]. Little data are available on hepatitis prevalence in Bangladesh specially in the context of Chattogram, southern part of Bangladesh [7]. So, present study is aimed to observe the frequency of different hepatotropic viruses causing hepatitis in our context.

## Methods

Present cross sectional observational study was conducted in a tertiary care hospital of Chattogram during a one year study periods from July 2022 to June 2023 among 114 patients. Patients with clinical jaundice or serum bilirubin <2 mg/dl with clinical diagnosis of hepatitis were the study subjects. After explanation of the study procedure informed written consent was taken from all patients to be included in the study. After getting the clinical data on a case record form, 5 ml of venous blood was collected to test the presence of hepatotropic viral markers like HAV, HBV, HCV and HEV. Testing was done by immunochromatographic(ICT) methods or Enzyme Linked Immunosorbent Assay(ELISA) for using standard testing kits available in the testing labs, CMOSHMC. Patients those were positive for HBV were undergone HIV testing also by ICT. All testing were done in clinical pathology and microbiology department of Chattogram Maa O Shishu Hospital and data were included in the Microsoft Excel for further analysis. Ethical clearance was obtained from the ERB of CMOSHMC to conduct the study. Funding of the study was provided by the CMOSHMC research fund from the yearly allocation to move research further forward.

## Results

Table 1 showing gender and age distributions of study patients where male were 73(64%) and female were 41(36%), male to female ratio was 1: 0.56. Maximum were in the age range of <20 years and 21- 30 years which were 46(40.4%) and 44(38.6%) respectively. Age group >60 years were 3(2.6%) only.

**Table 1:**
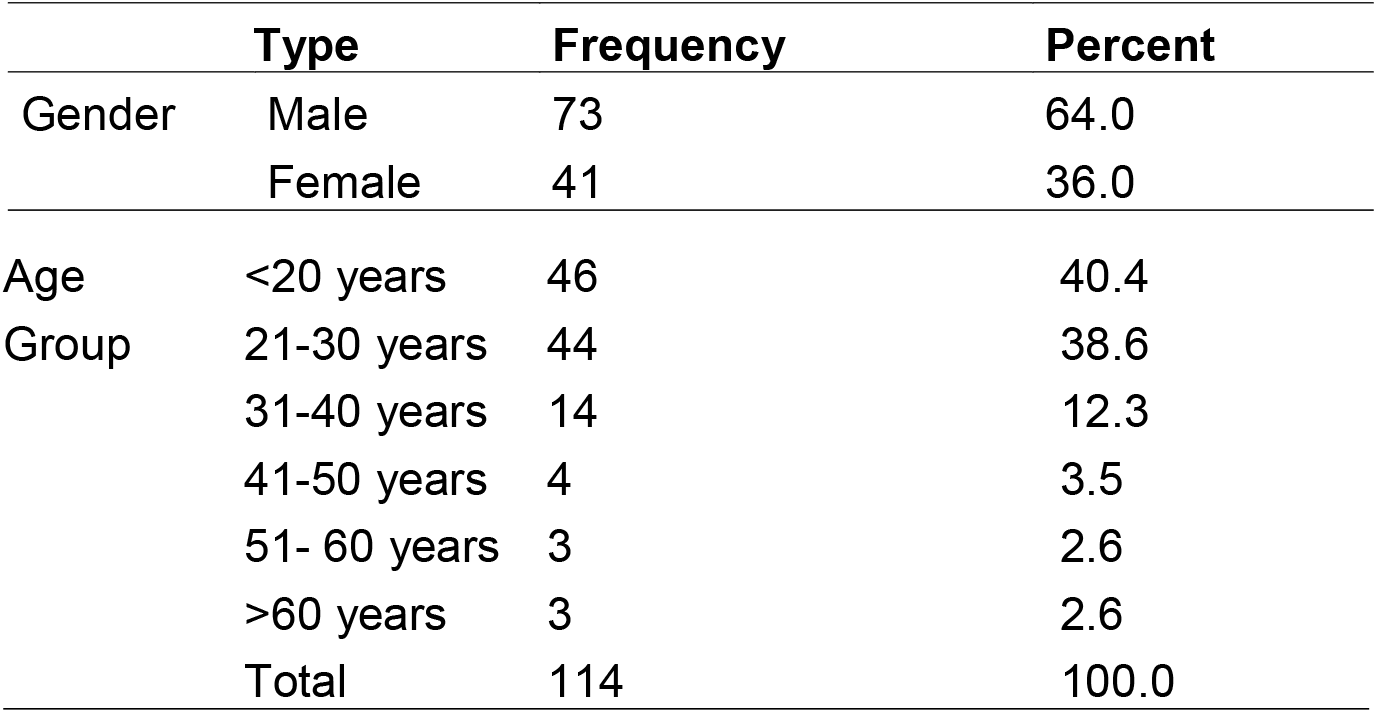
Age and gender distributions of the study patients.

Table 2 showing different clinical features of study patients where 109(95.6%) had jaundice and 89(78.1%) had fever and 74(64.9%) had nausea and these were three dominant clinical features. Regarding other features vomiting 7(6.1%), abdominal pain 66(57.9%), headache 4(3.5%), anorexia 30(24.4%) diarrhea 10(8.8%), fatigue 20(17.5%) and H/O liver disease were 8(7.0%)

**Table 2:** Clinical features of the patients.

| **Clinical features** | **Number** | **Percent** |
| --- | --- | --- |
| Fever | 89 | 78.1 |
| Nausea | 74 | 64.9 |
| Vomiting | 7 | 6.1 |
| Jaundice | 109 | 95.6 |
| Abdominal pain | 66 | 57.9 |
| Headache | 4 | 3.5 |
| Anorexia | 30 | 24.4 |
| Diarrhoea | 10 | 8.8 |
| Fatigue | 20 | 17.5 |
| H/O liver disease | 8 | 7.0 |

Table 3 showing frequency of HAV, HBV, HCV and HEV infection were 43(37.7%), 9(7.9%), 1(0.9%) and 26(22.8%) respectively. There were viral coinfections with (HAV+HEV), (HAV+HBV) and (HBV + HEV) and those were 8(7.0%), 4(3.5%) and 3(2.6%) respectively. Among all, 20(17.5%) cases were tested negative for hepatotropic viruses. Again 6(5.3%) had Anti HBc positivity and 2(1.8%) cases were found coinfected with HIV.

**Table 3:** Virologic patterns of hepatitis(N-114)

| **Types of diseases** | **Frequency** | **Percent** |
| --- | --- | --- |
| HAV | 43 | 37.7 |
| HBV | 9 | 7.9 |
| HCV | 1 | .9 |
| HEV | 26 | 22.8 |
| Unknown | 20 | 17.5 |
| HAV + HEV | 8 | 7.0 |
| HAV + HBV | 4 | 3.5 |
| HBV + HEV | 3 | 2.6 |
| Anti HBc Positivity | 6 | 5.3 |
| HIV coinfection | 2 | 1.8 |

## Discussion

In the present study, among 114 hepatitis cases analyzed, frequency of HAV, HBV, HCV and HEV infection were 43(37.7%), 9(7.9%), 1(0.9%) and 26(22.8%) respectively. Limited data are available on hepatitis prevalence in Bangladesh specially from southern part of Chittagong[7].But according to WHO, Bangladesh is one of the countries or areas with moderate to high risk ofhepatitis A and hepatitis B [7]. Bangladesh is considered to be a country wherehepatitis A infection is hyperendemic with 100% of children ≤ 6 years of age exposed and immuneto HAV[8] As a South East Asian country Bangladesh is considered endemic forhepatitis B virus (HBV) infection [9]. It was found that HEV carries alow mortality of 0.5-4% among the general populations but this figure approaches >75% in developing countries, such asBangladesh, in the second/third trimester of pregnancy, and in patients with fulminant hepaticFailure[10]. In Bangladesh,information about prevalence of HBV infection is scarce, and there is no available data on HDVinfection[11].In a study done among the HCWs, HBsAg positivity was found to be 8% in Bangladesh.[3]. A study conducted in Dhaka,Bangladesh by Ashraf *et al*.[12] found that the HBsAgprevalence of 6.5% among the common population 3%among healthy adults and children and 3.5% amongpregnant women; however, a much lower rate (0.8%)was observed among schoolchildren.

Among the HBV positive patients, two cases were also positive for HIV. Since both the hepatitis B virus and the HIV virusshare similar transmission routes, it isnot surprising that there is a high frequency of coinfection[13]. HBV infection is itself a dynamic disease and coinfectionwith HIV considerably complicates itsdiagnosis and management because the more rapiddevelopment of circulated HBV virus in the body.But co-infection with HBV is a preventable cause ofchronic liver disease among HIV-infected patients.[14,15]There were viral coinfections found in our study where (HAV+HEV), (HAV+HBV) and (HBV + HEV) and it were 8(7.0%), 4(3.5%) and 3(2.6%) respectively. HAV and HEV shares common routes of transmission[1] but coinfection with HBV with those may be incidental findings as routes of HBV transmission is not as same as the HAV or HEV. It needs further study in future.

Gender distributions revealed, malewere 73(64%) and female were 41(36%), male to female ratio was 1: 0.56. More hepatitis patients were in the age range of <20 years and 21- 30 years which were 46(40.4%) and 44(38.6%) respectively. In an earlier study regarding age range found more hepatitis were aggregated in younger age groups.[16]

Evaluations of different clinical features of study patients, 109(95.6%) had jaundice and 89(78.1%) had fever and 74(64.9%) had nausea and these were three dominant clinical features. Regarding others features vomiting 7(6.1%), abdominal pain 66(57.9%), headache 4(3.5%), anorexia 30(24.4%) diarrhea 10(8.8%), fatigue 20(17.5%) and H/O liver disease were 8(7.0%). In a study done byBattaet al.[17]found the sign and symptoms developed by the hepatitisaffected patients were classical sign and symptoms. Theyfound Jaundice (28.57%) and weight loss (28.57%) were mostcommon with loss of appetite (10.71%). It was an interestingobservation that 5.71 % of patients reported to have itchy skin.Itchiness is developed in particularly hepatitis A patients as aresult of cholestasis. So in the present study setting these are some common history andexamination findings among the patients of hepatitis and theseare as expected.[1] A significant number of patients of hepatitis were found virologically negative. So possibilities of other non hepatotropic viruses or different causes of hepatitis may be related with this findings and it needs further exploration and it may be an interesting topic of future research.

## Conclusion

HAV and HEV infections are the most common causes of acute hepatitisin Bangladesh as also found in our study. However, the real magnitude of HAV and HEV infections and HAV andHEVrelated acute hepatitis is under-recognized in Bangladesh, requiring further attention. HBV and HCV are two further threats for chronic hepatitis though former one is highly prevailing then the later in our country and Chattogram. Coinfection with HBV and HIV needs further attention.

## Data Availability

All data produced in the present study are available upon reasonable request to the authors

